# Running related biomechanical risk factors for overuse injuries in distance runners: A systematic review considering injury specificity and the potentials for future research

**DOI:** 10.1101/2021.07.23.21261034

**Authors:** Steffen Willwacher, Markus Kurz, Johanna Robbin, Matthias Thelen, Joseph Hamill, Luke Kelly, Patrick Mai

## Abstract

**Objective:** To identify and evaluate the evidence of the most relevant running-related risk factors (RRRFs) for running-related overuse injuries (ROIs) and to suggest future research directions.

**Design:** Systematic review considering prospective and retrospective studies. (PROSPERO_ID: 236832)

**Data sources:** Pubmed. Connected Papers. The search was performed in February 2021.

**Eligibility criteria:** English language. Studies on participants whose primary sport is running addressing the risk for the seven most common ROIs and at least one kinematic, kinetic (including pressure measurements), or electromyographic RRRF. An RRRF needed to be identified in at least one prospective or two retrospective studies.

**Results:** Sixty-two articles fulfilled our eligibility criteria. Levels of evidence for specific ROIs ranged from conflicting to moderate evidence. Running populations and methods applied varied considerably between studies. While some RRRFs appeared for several ROIs, most RRRFs were specific for a particular ROI. The biomechanical measurements performed in many studies would have allowed for consideration of many more RRRFs than have been reported, highlighting a potential for more effective data usage in the future.

**Conclusion:** This study offers a comprehensive overview of RRRFs for the most common ROIs, which might serve as a starting point to develop ROI-specific risk profiles of individual runners. Future work should use macroscopic (big data) approaches involving long-term data collections in the real world and microscopic approaches involving precise stress calculations using recent developments in biomechanical modelling. However, consensus on data collection standards (including the quantification of workload and stress tolerance variables and the reporting of injuries) is warranted.

## Introduction

Overuse injuries in runners (ROIs) are widespread, with a reported overall incidence of 19.4 to 79.3% [1]. Depending on the type of runner, definitions of injury, and follow-up periods, running-related injury incidence rates range between 2.5 to 33.0 injuries per 1000 hours of running [2]. The origins of ROIs are complex [3,4] but principally result from an accumulation of repetitive stress applied on the body without sufficient rest for tissue remodelling resulting in a degenerative response [5]. The stress response is a function of both tissue characteristics (influenced by lifestyle and genetic factors) and stress application characteristics (e.g., amplitude, frequency, duration) [6]. However, the non-invasive determination of these stresses is challenging as is the measurement of stress accumulation in everyday life and sports [7]. To determine structure specific stresses, computational models need to integrate precise anatomical information (e.g., biological tissues’ properties and geometry) and the potential neuromuscular control strategy that governs force and power production [8].

Therefore, researchers and practitioners often try to predict injury risk based on less direct and less computational and information-expensive biomechanical parameters as surrogate variables to link running biomechanics and injury risk. Such biomechanical running-related risk factors (RRRFs) include kinematic and kinetic parameters derived from ground reaction force, pressure mapping, electromyographic, and motion capture data. Using this approach, runners at risk of an ROI can be identified, however, to prevent ROIs, further knowledge on cause-effect relationships is needed [9].

Within a framework of injury development [10,11], the most relevant RRRFs could serve as a source for the improvement of technical (e.g., running shoes or foot orthoses / insoles), training and/or feedback system interventions (e.g., in gait retraining or through “digital coaches” based on wearable sensor information) by targeting populations at risk. Research on RRRFs employs different research designs and populations. The wealth of information is challenging to oversee.

Therefore, the aim of this review article is: (1) to identify the most relevant RRRFs and evaluate their evidence concerning the most prevalent ROIs; and (2) to suggest future directions of research to improve the understanding of the relationship between running biomechanics and overuse injury development while considering the interplay between RRRFs, workload characteristics and individual, structure-specific stress tolerances.

## Methods

### Search strategy and risk factor extraction

The systematic review aimed to extract the evidence for RRRFs for the ROIs with the highest prevalence and incidence. Therefore, based on the work of Lopes et al. [12], we examined RRRFs for the following ROIs: Medial tibial stress syndrome (MTSS), Achilles tendinopathy (AT), plantar fasciitis (PF), patellar tendinopathy (PT), iliotibial band syndrome (ITBS), tibial stress fracture (TSF), hamstring tendinopathy (HT), and patello-femoral pain syndrome (PFPS). We followed the reporting items for Systematic Reviews and Meta-analyses (PRISMA) guidelines [13]. Before starting the literature review, we registered this study at PROSPERO (record ID 236832).

We scanned the Pubmed database for articles comparing the running biomechanics of injured and uninjured individuals for the seven most common ROIs. For each ROI, we used an injury-specific search string (for details, please refer to the Supplementary Digital Content (SDC)). In short, each search string comprised combinations of runn* (i.e., the main activity), the injury location (e.g., femur*), multiple keywords to characterise injury-specific physical complaints (e.g., risk OR tend* OR pain), and the study design (e.g., prospective OR retrospective). We used an additional combination of keywords to obtain original English articles involving human participants (SDC). The initial search for ITBS, MTSS, and HT took place on February 4, 2021. One day after (February 5, 2021), the search strings for AT, PT, PFPS, PF, TSF were applied. Search results, including titles and abstracts, were uploaded to the web interface of rayyan.qcri.org [14]. We then screened titles and abstracts of the articles using the following criteria:

Inclusion criteria:

- Studies in the English language
- Prospective or retrospective studies addressing at least one of the ROIs of interest and relating injury risk to at least one RRRF
- Studies considering kinematic, kinetic (including pressure measurements), or electromyographic RRRFs
- The primary sport of the investigated study sample was running

Exclusion criteria:

- No RRRF analysed
- Studies that sampled from populations where distance running was not the primary sport
- Studies addressing biomechanical risk factors during dynamic activities other than running (e.g., walking or stair climbing)
- Studies addressing anthropometric factors (e.g., leg alignment, foot posture index) or strength measurements (e.g., toe strength or hip abduction strength)
- Studies including military or physical education students due to the unknown effects of concurrent training
- Studies (obviously) publishing results from the same subject sample as in a previous publication of the same group
- Non-original articles (e.g., reviews or conference articles) or articles not written in English

Two review team members independently selected titles and abstracts of studies found through the search strategy for potentially relevant studies after applying the inclusion and exclusion criteria. The selection of appropriate studies was discussed between the team members, and in the case of disagreements, they were resolved through consultation with another member of the review team. Subsequently, full texts were screened based on the same exclusion and inclusion criteria.

Additional sources were identified through the reference list of the eligible articles from the initial search and a co-citation method using the bibliographic coupling concept (http://www.connectedpapers.com).

Data on study characteristics were extracted from all included articles by members of the review team. Discrepancies were identified and resolved through discussion (with another reviewer if necessary). This data extraction included publication details (author and year), general information on injury type, specific running population, sample size, data collection method, running speed and footwear used during testing, and biomechanical outcome variables. Furthermore, we determined whether potential risk factors found in other studies could have been calculated based on the reported data collection methods. We also collected data on participant characteristics (e.g., age, gender, height).

### Relevance criterion for considering running-related risk factors

We considered an RRRF relevant if at least one prospective study or two retrospective studies from independent data collections found a significantly different value of an RRRF for a specific ROI.

### Quality rating and risk of bias assessment

We followed the same procedure as in a previously published review [15] using selected components from the ‘Quality Index’ developed by Downs and Black [16]. The modified ‘Quality Index’ scale consists of 15 items. All points of the modified ‘Quality Index’ were summed to provide a quality score for each study. Studies scoring 11 or greater were considered to be of high quality, studies with scores of six to ten were considered to be of moderate quality and studies with scores of five or less were considered to be of low quality [17]. Outcomes were discussed in a team meeting, and discrepancies were resolved by consulting another reviewer.

Due to poor reliability observed in items addressing external validity in the complete Downs and Black Quality Index [16], we performed a separate risk of bias assessment using a 10-point checklist, previously described in a systematic review of RRIs [12]. Each item was rated with either 1, referring to a low risk of bias, or 0, referring to a high risk of bias. If certain items could not be categorised, we assigned them a value of 0. Overall, we summed up the ten items’ scores. When less than half of the maximum possible points (i.e., <= 5 of 10 possible points) were reached, we considered the study to have a high risk of bias.

To determine the strength of evidence of an RRRF for a specific ROI, we followed the same approach as a previous review focussing on the role of RRRFs for running injuries in general [18]. These authors used the following categories described in detail by van Tulder et al. [19]:

- Strong evidence: Consistent findings among three or more studies, including a minimum of two high-quality studies
- Moderate evidence: Consistent findings among two or more studies, including at least one high-quality study
- Limited evidence: Findings from at least one high-quality study or two low- or moderate-quality studies
- Very limited evidence Findings from one low- or moderate-quality study
- Inconsistent evidence: Inconsistent findings among multiple studies (e.g., one or multiple studies reported a significant result, while one or multiple studies reported no significant result)
- Conflicting evidence We defined conflicting as contradictory results between studies (e.g., one or multiple studies reported a significant result in one direction, while one or multiple studies reported a significant result in the other direction)
- No evidence Results were insignificant and derived from multiple studies regardless of quality.

## Results

After identification, screening, and applying the exclusion and inclusion criteria, 62 articles were included in the review (Fig. 1).

**Figure 1:**
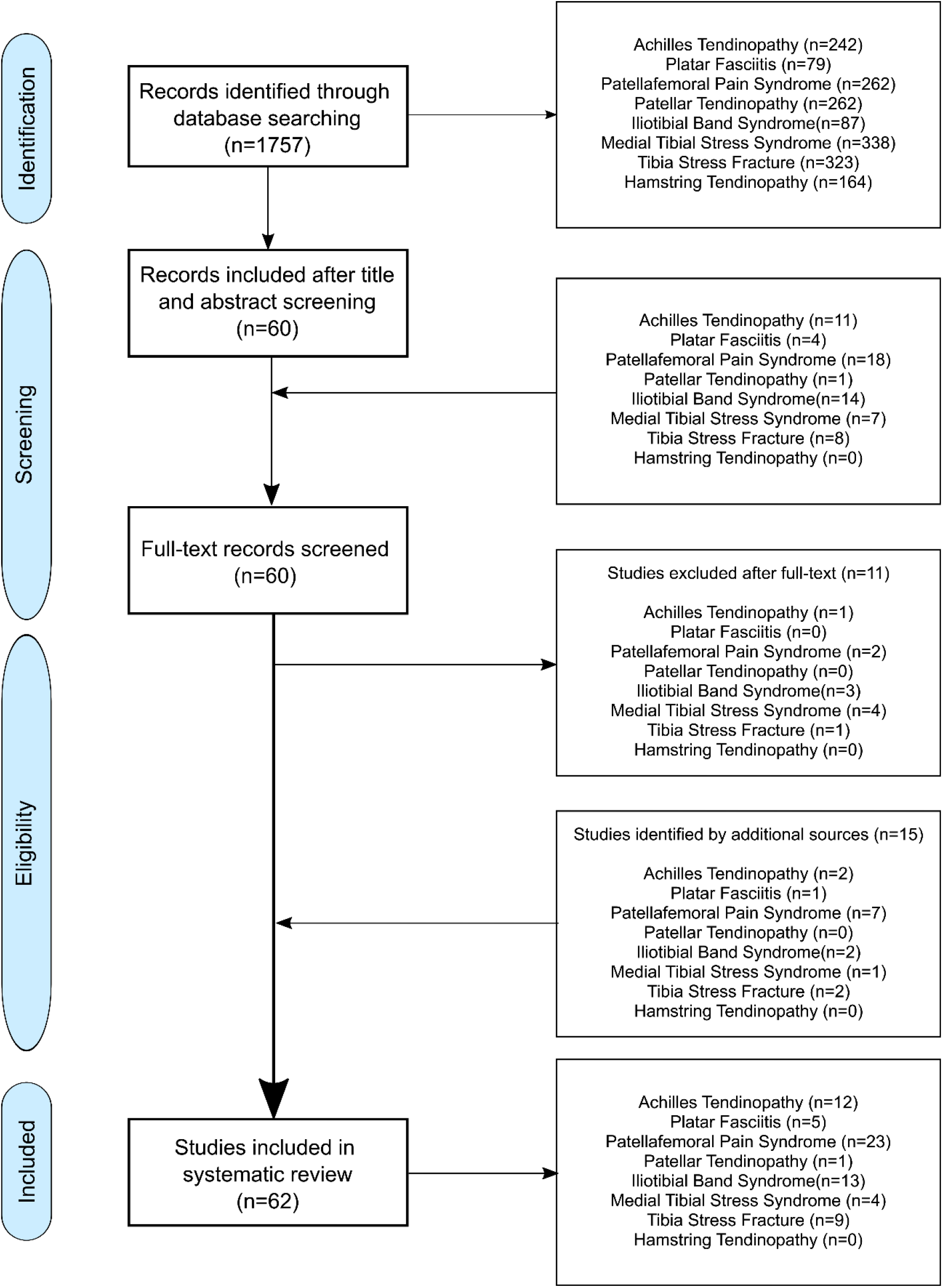
A FLOW-chart of the identification process. The numbers for articles per injury do not sum up to the total number of articles because some studies have addressed multiple running-related injuries.

In the following, we report the findings independently for each ROI considered. The findings are summarised graphically in Figure 2. Detailed results on study details, quality assessment and risk of bias rating can be found in the Supplementary Digital Contents (SDCs).

**Figure 2:**
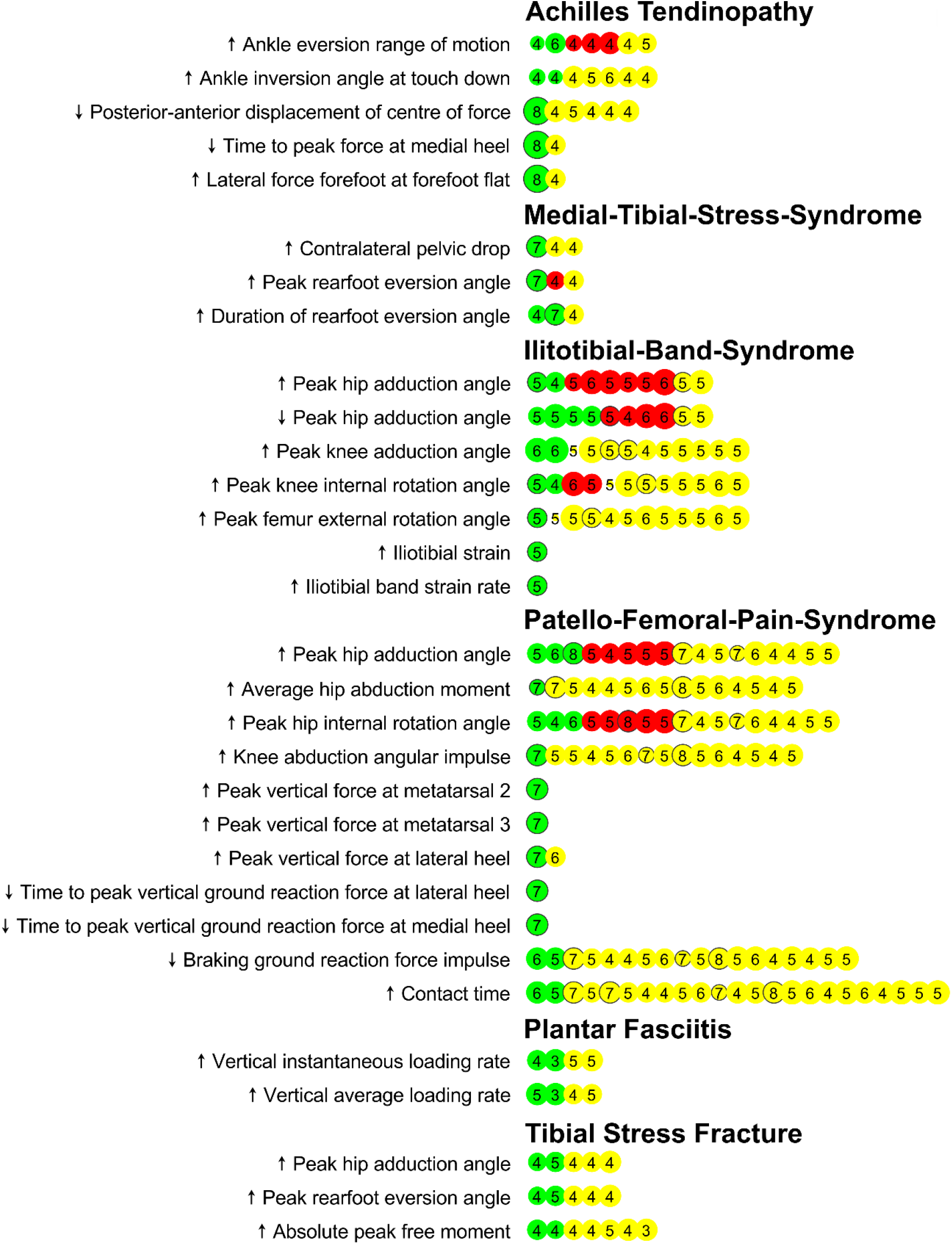
Graphical representation of the evidence associated with running-related risk factors that have passed our predefined relevance criterion (at least a significant difference in one prospective study or two retrospective studies). The green color represents a study that had found a significant difference between a group of injured runners compared to control. Red colors represent a study that could not find a significant difference between groups. Yellow colors represent studies that could have analysed a certain running-related risk factor based on the methodology they have used, but they did not consider the risk factor in their analysis. Black circles around dots indicate a prospective study design (no circles = retrospective study design). Dot size scales with Black & Down quality rating of the studies. The number in the dots is the Risk of Bias Score of the study.

### Achilles tendinopathy (AT)

We identified twelve studies (eleven retrospective, one prospective) that had analysed, in total, 115 different potential RRRFs for AT (SDC) through our systematic screening of the literature [20–31]. Out of these parameters, five RRRFs were identified in either two independent retrospective studies or one prospective study, following our predefined relevance criterion.

A high quality (B&D: 13) prospective study with low risk of bias (ROBS: 8) using a pressure plate found that novice runners who developed AT within a ten-week follow-up period showed three differences in their plantar pressure application during the stance phase compared to novice runners who remained injury-free. A reduced antero-posterior displacement of the centre of pressure during the stance phase, higher vertical forces applied through the lateral part of the foot at the instant of forefoot flat, and a reduced time to peak force at the medial heel.

Further, two RRRFs related to the motion of the ankle joint in the frontal plane (rearfoot inversion-eversion relative to the tibia) were identified. Two medium quality retrospective studies (B&D: 7-10), one with a high risk of bias (ROBS: 4), identified increased ankle range of motion from TD to maximum rearfoot eversion during the stance phase as RRRF [22,30]. Furthermore, more pronounced ankle inversion at initial contact with the ground was retrospectively identified as RRRF for AT by two medium quality studies with a high risk of bias (B&D: 7-8; ROBSs: 4). Many additional parameters differed between runners suffering from AT compared to runners who did not. For a complete list of all parameters for all ROIs, please refer to the SDC. However, these results were only found in single retrospective studies and did not follow our predefined quality criterion.

In summary, we identified limited evidence for a reduced anterior-posterior displacement of the centre of pressure, higher vertical forces applied through the lateral part of the foot at the instant of forefoot flat, and a reduced time to peak force at the medial heel during the stance phase as RRRFs for AT. We further found limited evidence for increased ankle inversion angle at initial contact and inconsistent evidence for ankle eversion range of motion from initial contact to peak rearfoot eversion during stance as RRRFs for AT (Fig. 3).

**Figure 3:**
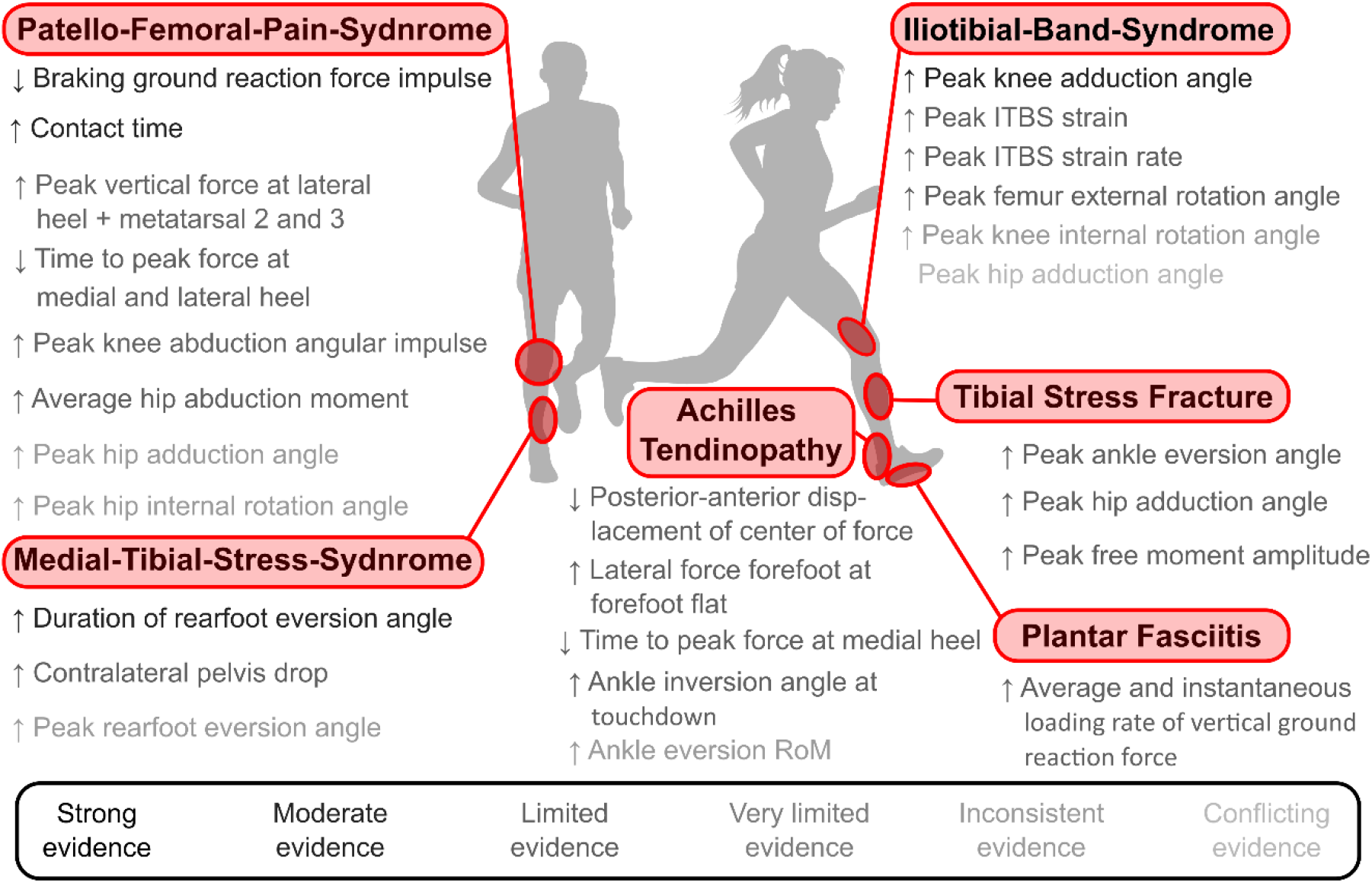
Overview of the evidence associated with running-related risk factors that have passed our predefined relevance criterion (at least a significant difference in one prospective study or two retrospective studies).

### Medial tibial stress syndrome (MTSS)

Our search resulted in four (one prospective, three retrospective studies) studies addressing RRRFs for MTSS [20,32–34]. In these studies, 23 individual RRRFs were investigated (SDC). However, only three RRRFs matched our relevance criterion.

In a high quality (B&D: 11; ROBS: 7) prospective study, competitive runners (NCAA Division 1) developing MTSS during a two-year follow-up period ran with greater peak rearfoot eversion relative to the tibia, and their ankle joints remained in an everted position for a longer time during the stance phase compared to runners not suffering from MTSS [32]. Furthermore, in the same study, runners developing MTSS had a higher contralateral peak pelvis drop during the stance phase compared to runners not suffering from MTSS [32]. The finding that runners with MTSS spend more time in eversion during stance was replicated in a moderate quality retrospective study (B&D: 9; ROBS: 4) [20]. However, the finding that peak eversion is a risk factor for MTSS was not replicated in this study [20].

In summary, we found moderate evidence for eversion time during stance, inconsistent evidence for peak eversion, and limited evidence for peak contralateral pelvis drop during stance as RRRFs for MTSS (Fig. 3).

### Tibial stress fractures (TSF)

We identified nine retrospective studies addressing RRRFs for TSF [31,35–42]. These studies considered 41 individual RRRFs (SDC). Three RRRFs followed our predefined relevance criterion. Two moderate quality studies (B&D: 9-10; ROBS: 4-5) found higher peak ankle eversion during stance for runners with a history of TSF [40,42]. These same studies also reported greater peak hip adduction angles during stance for runners with a history of TSF compared to runners without a history of TFS [40,42]. Further, two moderate quality studies (B&D: 9-10; ROBS: 4) found higher peak amplitudes of the free moment applied to the ground in runners with a history of TSF [39,42].

In summary, we identified limited evidence for peak ankle eversion, peak hip adduction, and peak free moment amplitude as RRRF for TSF (Fig. 3).

### Plantar fasciitis (PF)

Our search resulted in five retrospective studies considering 46 potential RRRFs for PF [31,43– 46]. Two out of these parameters matched with our predefined relevance criterion. Runners with a PF history created higher instantaneous vertical loading rates of the ground reaction force in two retrospective studies [31,45]. One study was of high quality (B&D: 11) but also a high risk of bias (ROBS: 3) [31], while the other study was of moderate quality (B&D: 10) and also a high risk of bias (ROBS: 4) [45]. Further, two high-quality (B&D: 11; ROBS: 3-5) studies found that runners with PF history applied vertical forces at a higher average loading rate to the ground [31,43].

In summary, we found limited evidence for average and instantaneous vertical loading rates of the ground reaction force as RRRFs for PF (Fig. 3).

#### Iliotibial band syndrome (ITBS)

We found 13 studies (three prospective and ten retrospective) considering 71 potential RRRFs for ITBS [31,46–57]. Of these parameters, seven followed our relevance criterion. At the hip, conflicting evidence was found for the peak hip adduction angle. While one moderate quality retrospective study (B&D: 10; ROBS: 4) [53] and one moderate quality prospective study (B&D: 10; ROBS: 5%) [56] found significantly higher peak hip adduction angles in runners with ITBS, three moderate (B&D: 9-10; ROBS: 5) and two high quality (B&D: 12; ROBS: 4-5) retrospective studies found reduced peak hip adduction angles during the stance phase in runners with ITBS compared to non-injured runners (Fig. 2).

A moderate quality (B&D: 10; ROBS: 5) prospective study found higher peak external rotation during stance in runners who developed ITBS compared to their control group [56] (Fig. 2). At the knee, a moderate quality retrospective study (B&D: 10; ROBS: 4) [53] and one moderate quality prospective study (B&D: 10; ROBS: 5) [56] found significantly higher peak internal rotation angles during the stance phase in runners with ITBS compared to non-injured runners. Further, two high-quality retrospective studies (B&D: 12-13; ROBS: 6) reported significantly higher peak knee adduction angles in runners with compared to runners without a history of ITBS [49,52]. When applying a computer model which calculates the kinematics of the ITB, Hamill et al. [55] identified increased ITB strain and strain rates in runners with compared to runners without a history of ITBS (Fig. 2).

In summary, our systematic review established moderate evidence for increased peak knee adduction angle, limited evidence for increased ITB strain, increased ITB strain rate and increased peak femur external rotation. Further, we found inconsistent evidence for increased peak knee internal rotation angle and conflicting evidence for peak hip adduction as an RRRF for ITBS (Fig. 3).

#### Patello-femoral pain syndrome (PFPS)

Twenty-three studies (four prospective and 19 retrospective) were included in the systematic review [31,58–61,61–78]. These studies analysed in total 114 potential RRRFs (SDC). Of these, eleven RRRFs matched our predefined relevance criterion. Longer contact time was identified as an RRRF for PFPS in two high-quality (B&D: 11) retrospective studies with a low risk of bias (ROBS: 5-6)[71,74]. Several plantar pressure-related variables were identified by one high-quality (B&D: 11) prospective study with a low risk of bias (ROBS: 7) [70]. These were an increased peak vertical force at the lateral heel (i.e., initial contact), as well as at the 2^nd^ and 3^rd^ metatarsal heads. Further, a reduced time to peak force at the medial and lateral heel were found. Two retrospective studies (B&D: 10-11; ROBS: 5-6) related a reduced braking impulse of the horizontal ground reaction force with an increased risk for PFPS [71,74] (Fig. 2).

One high-quality (B&D: 11) prospective study with low risk of bias (ROBS: 70%) found greater internal knee abduction angular impulses in runners developing PFPS compared to non-injured controls [67] (Fig. 2). At the hip, higher peak adduction angles were identified as risk factors by one high quality prospective study with low risk of bias (D&B: 11; ROBS: 8) [62] and two retrospective studies (D&B: 10; ROBS: 5-6) [69,72]. Further, three moderate quality retrospective studies (B&D: 10, ROBS: 4-6) suggested that an increased peak hip internal rotation angle was associated with PFPS [60,69,72]. A moderate quality prospective study with low risk of bias (B&D: 8; ROBS: 7) found increased average internal hip abduction moments in runners who developed PFPS compared to runners who did not [63] (Fig. 2).

In summary, we found limited evidence for above mentioned plantar pressure-related parameters, increased internal knee abduction angular impulse, and increased average hip internal abduction moments during stance. Further, moderate evidence for reduced braking impulse of the ground reaction force and longer ground contact times, and inconsistent evidence for increased peak hip adduction and internal rotation angles during stance were found (Fig. 3).

#### Patellar and hamstring tendinopathy (PHT)

A moderate quality study with a high risk of bias (B&D: 7; ROBS: 3) analysed 42 potential RRRFs for PHT [79]. However, since this was the only study, our predefined relevance criterion was not met. We could not identify a study focussing on RRRFs for hamstring tendinopathy.

## Discussion

This systematic review aimed to extract the evidence for RRRFs for specific ROISs from the existing literature. While there are several important previous reviews on the role of RRRFs for the development of running-related injuries, our work adds several relevant pieces to the complex puzzle of ROI development. It is the first systematic review that focuses on RRRFs for the most prevalent ROIs while using the same inclusion and exclusion criteria for all considered injuries. Previous reviews either did not report overuse injuries for specific types of injuries [18] or focus on a single overuse injury [15,80–88]. Further, some reviews did only focus on prospective studies [18]. While these studies are superior in their strength of evidence to retrospective studies, the majority of research on RRRFs for ROIs have used retrospective designs. By applying our relevance criterion (potential RRRFs identified from at least two retrospective studies or one prospective study), we acknowledged the superior evidence of the prospective study design while at the same time including insight gained from retrospective studies.

A list of relevant RRRFs can be used to establish an individual risk profile of an individual’s running biomechanics. Based on this risk profile, individualised footwear could be developed, or footwear might be reconsidered to change the running biomechanics towards a less risky profile. Considering specific injuries is significant progress for injury profiling since running shoes can be designed to shift loading between musculoskeletal structures in the lower extremity and hence specifically address injury-specific risk factors [5]. A running injury risk profile can also inform prevention training programs to strengthen biological tissues at risk or help to develop feedback tools that facilitate running gait retraining towards a less pronounced risk profile. More sophisticated, while at the same time easy-to-use (maybe integrated with running apparel), tools for gait retraining may be available in the future [89].

Since a previous injury is an essential non-running related injury risk factor [90], it seems logical that individualised running shoe design, prevention training protocols, or running gait feedback tools should weigh specific RRRFs based on injury history.

Further, it has to be clearly mentioned that RRRFs should be viewed in interaction with workload characteristics of training protocols and individual, structure specific load tolerance levels. This means that an individual risk profile based on RRRFs and potentially other risk factors can only inform interventions for runners if workload characteristics and stress tolerance levels are considered. Clearly more research is needed to improve our understanding on these different aspects of ROI development.

### Limitations

One of the strengths of our review is that it systematically assessed the current evidence for biomechanical RRRFs specifically for the most prevalent ROIs using the same methodology. Further, it includes work from both, propsepective and retrospective studies while acknowledging the superior level of evidence of prospective studies by applying a relevance criterion for retrospective study inclusion. Finally it highlights the potential for a more effective data usage by identying how additional RRRFs could have been analysed already in previous studies in the past.

Despite the several strengths of this work, we need to also highlight several limitations: Due to the lack of results reported or analysed in the considered studies, we could not differentiate our findings for different groups of runners. Since different groups of runners likely vary in their structure specific stress tolerance levels and their adaptations we recommend that future work on RRRFs should always report as many details of the running population as possible.

Further, different studies used heterogeneous definitions of injury, the definition of types of runners (e.g., competitive vs. recreational), and outcome measures in the included full-text articles challenged comparison across studies. Also, most studies did not consider running volume in their assessment of injury risk between groups (e.g., Incidence per 1000h of running [2]) or tried to quantify workload characteristics by other means.

### Outlook

Based on the findings of this review and when considering recent injury development frameworks, we propose the following directions for future research. These directions can be broadly categorised by either using larger datasets with lower data precision or smaller datasets with higher precision.

The big data macroscopic approach can leverage the recent developments in wearable sensor technology and artificial intelligence. At this time, running movement data can be captured during every training session and uploaded to large databases. The insight gained from the big data approach relies on the ability to determine relevant features (i.e., functional or discrete features related to injury risk) from these sensor signals, with the assistance of artificial intelligence. The parameters identified from this review can serve as a starting point for such a data exploration. Future studies need to explore the potential to identify new RRRFs using wearable, inexpensive sensor technology outside the laboratory setting. Further, tools to collect and store data on large scales while using user feedback to label the occurrence of running-related pain or injuries will allow further insight by considering not only single data collection sessions but, in principle, the entire training history of an individual (e.g., changes associated with fatigue) [91–93]. International research collaborations that use the same methodology for data capturing and labeling seem ideally suited to solve this task.

The current review findings also highlight that there already exists a “data treasure” from research performed over the last few decades which could be reanalysed by considering recent findings. Figure 2 highlights that several RRRFs could be analysed from existing datasets. A statistical reanalysis of the differences between injured and non-injured groups for these RRRFs from previous studies would help paint a clearer picture of the relationship between RRRFs and specific running injuries.

The small data microscopic approach relies on improvements in biomechanical modeling approaches that can improve our understanding of how running biomechanics are linked to the stress of the tissues involved in ROIs. Here, the combination of individualised musculoskeletal models with, for example, finite element models of the tissues under consideration seems to offer enormous potential for not only improved targeting of runner populations at risk but also increased understanding of cause effect relationships in ROI development. Single subjects study designs applying very detailed modelling techniques might further improve our understanding of injury development since the etiology of an injury is not the same for all patients diagnosed with the same injury. However, currently, these techniques are time-consuming and rely on many assumptions that challenge the validity of the calculated stress characteristics. Therefore, the discipline of biomechanics should also target a more efficient yet precise quantification of input variables for these model calculations.

Independent of the scale and precision, running-related risk factors should not be considered in isolation but need to be analysed while considering the complex framework of running injury aetiology [11]. Therefore, the interplay of RRRFs with other modifiable and non-modifiable risk factors, workload characteristics [7], and estimators of structure-specific stress tolerance levels should be included in data collections and statistical analyses. A consensus on the minimum number and type of such framework variables for running injury research seems urgently needed to face this challenge.

In summary, this is the first systematic review that summarises the evidence for RRRFs for specific ROIs using the same search strategy and exclusion and inclusion criteria. We hope that this work can serve as the basis to identify runners at risk for specific ROIs and and from this basis improve decisions on footwear design or use, training and rehabilitation programs, and sensor-based devices to monitor and improve individual running biomechanics.

## Supporting information

Supplementary Digital Content 1

Supplementary Digital Content 2

Supplementary Digital Content 3

## Data Availability

The data undrlying this review are available from the Supplementary digital contents.

## Acknowledgements

There are no competing interests to declare. The preparation of this review was not financially supported.

