## Supplementary Digital Content 1 for "Running related biomechanical risk factors for overuse injuries in distance runners: A systematic review considering injury specificity and the potentials for future research"

**Achilles tendinopathy**

Date of Access: 5.2.2021: N = 242

(runn*) AND (injur* OR risk OR factor* OR tend* OR syndrome OR stress* OR fasc* OR pain) AND (prospective OR retrospective OR cross* OR long* OR follow* OR case* OR cohort*) AND (achil*) NOT (“addresses”[Publication Type] OR “bibliography”[Publication Type] OR “biography”[Publication Type] OR “case reports”[Publication Type] OR “clinical conference”[Publication Type] OR “comment”[Publication Type] OR “congresses”[Publication Type] OR “dictionary”[Publication Type] OR “directory”[Publication Type] OR “editorial”[Publication Type] OR “festschrift”[Publication Type] OR “government publications”[Publication Type] OR “interview”[Publication Type] OR “lectures”[Publication Type] OR “legal cases”[Publication Type] OR “legislation”[Publication Type] OR “letter”[Publication Type] OR “news”[Publication

Type] OR “newspaper article”[Publication Type] OR “retracted publication”[Publication Type] OR “retraction of publication”[Publication Type] OR “review”[Publication Type] OR “review literature”[Publication Type] OR “review of reported cases”[Publication Type] OR “review, academic”[Publication Type] OR “review, multicase”[Publication Type] OR “review, tutorial”[Publication Type] OR “scientific integrity review”[Publication Type] OR “technical report”[Publication Type] OR “twin study”[Publication Type] OR “validation studies”[Publication Type]) AND “Humans”[Species]

**iliotibial band friction**

Date of Access: 4.2.2021: N = 87

(runn*) AND (injur* OR risk OR factor* OR tend* OR syndrome OR stress* OR fasc* OR pain OR friction) AND (prospective OR retrospective OR cross* OR long* OR follow* OR case*) AND (ilio*) NOT (“addresses”[Publication Type] OR “bibliography”[Publication Type] OR “biography”[Publication Type] OR “case reports”[Publication Type] OR “clinical conference”[Publication Type] OR “comment”[Publication Type] OR “congresses”[Publication Type] OR “dictionary”[Publication Type] OR “directory”[Publication Type] OR “editorial”[Publication Type] OR “festschrift”[Publication Type] OR “government publications”[Publication Type] OR “interview”[Publication Type] OR “lectures”[Publication Type] OR “legal cases”[Publication Type] OR “legislation”[Publication Type] OR “letter”[Publication Type] OR “news”[Publication Type] OR “newspaper article”[Publication Type] OR “retracted publication”[Publication Type] OR “retraction of publication”[Publication Type] OR “review”[Publication Type] OR “review literature”[Publication Type] OR “review of reported cases”[Publication Type] OR “review, academic”[Publication Type] OR “review, multicase”[Publication Type] OR “review, tutorial”[Publication Type] OR “scientific integrity review”[Publication Type] OR “technical report”[Publication Type] OR “twin study”[Publication Type] OR “validation studies”[Publication Type]) AND “Humans”[Species]

**Medial tibial stress syndrome**

Date of Access: 4.2.2021: N = 338

(runn*) AND (injur* OR risk OR factor* OR tend* OR syndrome OR stress* OR fasc* OR pain OR femu* OR frac* OR medial) AND (prospective OR retrospective OR cross* OR long* OR follow* OR cohort) AND (tibia*) NOT (“addresses”[Publication Type] OR “bibliography”[Publication Type] OR “biography”[Publication Type] OR “case reports”[Publication Type] OR “clinical conference”[Publication Type] OR “comment”[Publication Type] OR “congresses”[Publication Type] OR “dictionary”[Publication Type] OR “directory”[Publication Type] OR “editorial”[Publication Type] OR “festschrift”[Publication Type] OR “government publications”[Publication Type] OR “interview”[Publication Type] OR “lectures”[Publication Type] OR “legal cases”[Publication Type] OR “legislation”[Publication Type] OR “letter”[Publication Type] OR “news”[Publication Type] OR “newspaper article”[Publication Type] OR “retracted publication”[Publication Type] OR “retraction of publication”[Publication Type] OR “review”[Publication Type] OR “review literature”[Publication Type] OR “review of reported cases”[Publication Type] OR “review, academic”[Publication Type] OR “review, multicase”[Publication Type] OR “review, tutorial”[Publication Type] OR “scientific integrity review”[Publication Type] OR “technical report”[Publication Type] OR “twin study”[Publication Type] OR “validation studies”[Publication Type]) AND “Humans”[Species]

**Tibia stress fracture**

Date of Access: 5.2.2021: N = 323

(runn*) AND (injur* OR risk OR factor* OR tend* OR syndrome OR stress* OR fasc* OR pain OR femu* OR frac*) AND (prospective OR retrospective OR cross* OR long* OR follow* OR cohort) AND (tibia*) NOT (“addresses”[Publication Type] OR “bibliography”[Publication Type] OR “biography”[Publication Type] OR “case reports”[Publication Type] OR “clinical conference”[Publication Type] OR “comment”[Publication Type] OR “congresses”[Publication Type] OR “dictionary”[Publication Type] OR “directory”[Publication Type] OR “editorial”[Publication Type] OR “festschrift”[Publication Type] OR “government publications”[Publication Type] OR “interview”[Publication Type] OR “lectures”[Publication Type] OR “legal cases”[Publication Type] OR “legislation”[Publication Type] OR “letter”[Publication Type] OR “news”[Publication

Type] OR “newspaper article”[Publication Type] OR “retracted publication”[Publication Type] OR “retraction of publication”[Publication Type] OR “review”[Publication Type] OR “review literature”[Publication Type] OR “review of reported cases”[Publication Type] OR “review, academic”[Publication Type] OR “review, multicase”[Publication Type] OR “review, tutorial”[Publication Type] OR “scientific integrity review”[Publication Type] OR “technical report”[Publication Type] OR “twin study”[Publication Type] OR “validation studies”[Publication Type]) AND “Humans”[Species]

**Plantar fasciitis**

Date of Access: 5.2.2021: N = 79

(runn*) AND (injur* OR risk OR factor* OR tend* OR syndrome OR stress* OR fasc* OR pain) AND (prospective OR retrospective OR cross* OR long* OR follow* OR case* OR cohort*) AND ((plant* AND fasc*) OR (heel* AND spur*)) NOT (“addresses”[Publication Type] OR “bibliography”[Publication Type] OR “biography”[Publication Type] OR “case reports”[Publication Type] OR “clinical conference”[Publication Type] OR “comment”[Publication Type] OR “congresses”[Publication Type] OR “dictionary”[Publication Type] OR “directory”[Publication Type] OR “editorial”[Publication Type] OR “festschrift”[Publication Type] OR “government publications”[Publication Type] OR “interview”[Publication Type] OR “lectures”[Publication Type] OR “legal cases”[Publication Type] OR “legislation”[Publication Type] OR “letter”[Publication Type] OR “news”[Publication Type] OR “newspaper article”[Publication Type] OR “retracted publication”[Publication Type] OR “retraction of publication”[Publication Type] OR “review”[Publication Type] OR “review literature”[Publication Type] OR “review of reported cases”[Publication Type] OR “review, academic”[Publication Type] OR “review, multicase”[Publication Type] OR “review, tutorial”[Publication Type] OR “scientific integrity review”[Publication Type] OR “technical report”[Publication Type] OR “twin study”[Publication Type] OR “validation studies”[Publication Type]) AND “Humans”[Species]

**Patellofemoral pain syndrome**

Date of Access: am 5.2.2021: N = 262

(runn*) AND (injur* OR risk OR factor* OR tend* OR syndrome OR stress* OR fasc* OR pain OR femo* OR femu*) AND (prospective OR retrospective OR cross* OR long* OR follow* OR cohort) AND (patel*) NOT (“addresses”[Publication Type] OR “bibliography”[Publication Type] OR “biography”[Publication Type] OR “case reports”[Publication Type] OR “clinical conference”[Publication Type] OR “comment”[Publication Type] OR “congresses”[Publication Type] OR “dictionary”[Publication Type] OR “directory”[Publication Type] OR “editorial”[Publication Type] OR “festschrift”[Publication Type] OR “government publications”[Publication Type] OR “interview”[Publication Type] OR “lectures”[Publication Type] OR “legal cases”[Publication Type] OR “legislation”[Publication Type] OR “letter”[Publication Type] OR “news”[Publication Type] OR “newspaper article”[Publication Type] OR “retracted publication”[Publication Type] OR “retraction of publication”[Publication Type] OR “review”[Publication Type] OR “review literature”[Publication Type] OR “review of reported cases”[Publication Type] OR “review, academic”[Publication Type] OR “review, multicase”[Publication Type] OR “review, tutorial”[Publication Type] OR “scientific integrity review”[Publication Type] OR “technical report”[Publication Type] OR “twin study”[Publication Type] OR “validation studies”[Publication Type]) AND “Humans”[Species]

**Patellar tendinopathy Jumpers knee**

Date of Access: 5.2.2021: N = 262

(runn*) AND (injur* OR risk OR factor* OR tend* OR syndrome OR stress* OR fasc* OR pain OR femo* OR femu*) AND (prospective OR retrospective OR cross* OR long* OR follow* OR cohort) AND (patel* OR (jump* AND knee*)) NOT (“addresses”[Publication Type] OR “bibliography”[Publication Type] OR “biography”[Publication Type] OR “case reports”[Publication Type] OR “clinical conference”[Publication Type] OR “comment”[Publication Type] OR “congresses”[Publication Type] OR “dictionary”[Publication Type] OR “directory”[Publication Type] OR “editorial”[Publication Type] OR “festschrift”[Publication Type] OR “government publications”[Publication Type] OR “interview”[Publication Type] OR “lectures”[Publication Type] OR “legal cases”[Publication Type] OR “legislation”[Publication Type] OR “letter”[Publication Type] OR “news”[Publication Type] OR “newspaper article”[Publication Type] OR “retracted publication”[Publication Type] OR “retraction of publication”[Publication Type] OR “review”[Publication Type] OR “review literature”[Publication Type] OR “review of reported cases”[Publication Type] OR “review, academic”[Publication Type] OR “review, multicase”[Publication Type] OR “review, tutorial”[Publication Type] OR “scientific integrity review”[Publication Type] OR “technical report”[Publication Type] OR “twin study”[Publication Type] OR “validation studies”[Publication Type]) AND “Humans”[Species]

**Hamstring tendinopathy**

Date of Access: 5.2.2021: N = 164

(runn*) AND (injur* OR risk OR factor* OR tend* OR syndrome OR stress* OR fasc* OR pain) AND (prospective OR retrospective OR cross* OR long* OR follow* OR case* OR cohort) AND (hamstring*) NOT ("addresses"[Publication Type] OR "bibliography"[Publication Type] OR "biography"[Publication Type] OR "case reports"[Publication Type] OR "clinical conference"[Publication Type] OR "comment"[Publication Type] OR "congresses"[Publication Type] OR "dictionary"[Publication Type] OR "directory"[Publication Type] OR "editorial"[Publication Type] OR "festschrift"[Publication Type] OR "government publications"[Publication Type] OR "interview"[Publication Type] OR "lectures"[Publication Type] OR "legal cases"[Publication Type] OR "legislation"[Publication Type] OR "letter"[Publication Type] OR "news"[Publication Type] OR "newspaper article"[Publication Type] OR "retracted publication"[Publication Type] OR "retraction of publication"[Publication Type] OR "review"[Publication Type] OR "review literature"[Publication Type] OR "review of reported cases"[Publication Type] OR "review, academic"[Publication Type] OR "review, multicase"[Publication Type] OR "review, tutorial"[Publication Type] OR "scientific integrity review"[Publication Type] OR "technical report"[Publication Type] OR "twin study"[Publication Type] OR "validation studies"[Publication Type]) AND "Humans"[Species]
