## Supplementary Digital Content 2 for "Running related biomechanical risk factors for overuse injuries in distance runners: A systematic review considering injury specificity and the potentials for future research"

### Quality Assessment – Black & Down Scoring

#### Achilles tendinopathy

| | Prospective (P) or retrospective (R) study | Clear aim/hypothesis | Outcome measures clearly described | Patient characteristics clearly described | Confounding variables described | Main findings clearly described | Measures of random variability provided | Actual probability values reported | Participants asked to participate representative of entire population | Participants prepared to participate representative of entire population | Blinding of outcome measurer | Analysis completed was planned | Appropriate statistics | Valid and reliable outcome measures | Appropriate case-control matching | Adjustment made for confounding variables | Total |
|---|---|---|---|---|---|---|---|---|---|---|---|---|---|---|---|---|---|
| McCrory et al. 1999 | R | 1 | 1 | 0 | 1 | 1 | 1 | 0 | U | U | U | 1 | 1 | U | U | 1 | 8 |
| Donoghue et al. 2008 | R | 1 | 1 | 0 | ~~1~~ | 1 | 1 | 0 | 0 | 0 | U | 1 | 1 | U | U | 0 | 7 |
| Williams et al., 2008 | R | 1 | 1 | 1 | 1 | 1 | 1 | 1 | U | U | U | 1 | 1 | 1 | 1 | U | 11 |
| Azevedo et al., 2009 | R | 1 | 1 | 1 | 1 | 1 | 1 | 1 | U | U | U | 1 | 1 | 1 | U | U | 10 |
| Ryan et al. 2009 | R | 1 | 0 | 1 | 1 | 1 | 1 | 1 | U | U | U | 1 | 1 | U | 1 | 1 | 10 |
| van Ginckle et al. 2009 | P | 1 | 1 | 1 | 2 | 1 | 1 | 1 | 1 | U | 1 | 1 | 1 | U | 1 | 1 | 14 |
| Baur et al., 2011 | R | 1 | 1 | 1 | 1 | 1 | 1 | 1 | U | U | U | 1 | 1 | 1 | U | 0 | 10 |
| Wyndow et al. 2013 | R | 1 | 1 | 1 | 1 | 1 | 1 | 1 | U | U | U | 1 | 1 | 1 | U | 1 | 11 |
| Franettovich et al. 2014 | R | 1 | 1 | 1 | 1 | 1 | 1 | 1 | U | U | U | 1 | 1 | U | U | U | 9 |
| Creaby et al. 2016 | R | 1 | 1 | 1 | 1 | 1 | 1 | 1 | U | U | U | 1 | 1 | 1 | U | U | 10 |
| Becker et al. 2017 | R | 1 | 1 | 1 | 1 | 1 | 1 | 0 | 1 | U | U | 1 | 1 | 1 | 1 | U | 11 |
| Johnson et al. 2020 | R | 1 | 1 | 1 | 1 | 1 | 1 | 1 | U | U | 1 | 1 | 1 | 1 | U | U | 11 |

### Iliotibial band syndrome

| | Prospective (P) or retrospective (R) study | Clear aim/hypothesis | Outcome measures clearly described | Patient characteristics clearly described | Confounding variables described | Main findings clearly described | Measures of random variability provided | Actual probability values reported | Participants asked to participate representative of entire population | Participants prepared to participate representative of entire population | Blinding of outcome measurer | Analysis completed was planned | Appropriate statistics | Valid and reliable outcome measures | Appropriate case-control matching | Adjustment made for confounding variables | Total |
|---|---|---|---|---|---|---|---|---|---|---|---|---|---|---|---|---|---|
| Brown et al. 2019 | R | 1 | 1 | 1 | 1 | 1 | 1 | 1 | 1 | 0 | U | 1 | 1 | 1 | 0 | 1 | 12 |
| Baker et al. 2018 | R | 1 | 1 | 1 | 1 | 1 | 1 | 1 | 1 | U | U | 1 | 1 | 1 | 1 | 1 | 13 |
| Brown et al. 2016 | R | 1 | 1 | 1 | 1 | 1 | 1 | 1 | 0 | 0 | U | 1 | 1 | 1 | U | 0 | 10 |
| Foch et al. 2015 | R | 1 | 1 | 1 | 1 | 1 | 1 | 1 | 0 | 0 | U | 1 | 1 | 1 | U | 0 | 10 |
| Phinyomark et al., 2014 | R | 1 | 1 | 1 | 1 | 0 | 0 | 0 | 1 | U | U | 1 | 1 | 1 | 1 | 0 | 9 |
| Noehren et al. 2014 | R | 1 | 1 | 1 | 1 | 1 | 1 | 1 | 0 | 0 | U | 1 | 1 | 1 | 1 | 1 | 12 |
| Foch and Milner 2014 | R | 1 | 1 | 1 | 1 | 1 | 1 | 1 | 1 | 0 | U | 1 | 1 | 1 | 1 | 0 | 12 |
| Ferber et al. 2010 | R | 1 | 1 | 1 | 1 | 1 | 1 | 1 | 0 | 0 | U | 1 | 1 | 1 | U | 0 | 10 |
| Grau et al 2008 | R | 1 | 1 | 1 | 1 | 1 | 1 | 1 | 0 | 0 | U | 1 | 1 | 1 | 1 | 1 | 12 |
| Hamill et al. 2008 | P | 1 | 1 | 1 | 1 | 1 | 1 | 1 | 0 | 0 | U | 1 | 1 | 1 | U | 0 | 10 |
| Noehren et al. 2007 | P | 1 | 1 | 1 | 1 | 1 | 1 | 1 | 0 | 0 | U | 1 | 1 | 1 | U | 0 | 10 |
| Messier et al. 1995 | R | 1 | 1 | 1 | 1 | 1 | 1 | 1 | 1 | 1 | U | 1 | 1 | 1 | U | 1 | 13 |
| Johnson et al., 2020 | R | 1 | 1 | 1 | 1 | 1 | 1 | 1 | U | U | 1 | 1 | 1 | 1 | U | U | 11 |
| Grau et al 2011 | R | 1 | 1 | 1 | 1 | 1 | 1 | 0 | 0 | 0 | U | 1 | 1 | 1 | U | 0 | 9 |
| Messier et al 1988 | R | 1 | 0 | 0 | 0 | 0 | U | 0 | U | U | 0 | 1 | 1 | 0 | U | U | 3 |
| Millert et al. 2007 | R | 1 | 1 | 1 | 1 | 1 | 1 | 1 | U | U | U | 1 | 1 | 1 | 0 | U | 10 |

### Medial tibial stress syndrome

| | Prospective (P) or retrospective (R) study | Clear aim/hypothesis | Outcome measures clearly described | Patient characteristics clearly described | Confounding variables described | Main findings clearly described | Measures of random variability provided | Actual probability values reported | Participants asked to participate representative of entire population | Participants prepared to participate representative of entire population | Blinding of outcome measurer | Analysis completed was planned | Appropriate statistics | Valid and reliable outcome measures | Appropriate case-control matching | Adjustment made for confounding variables | Total |
|---|---|---|---|---|---|---|---|---|---|---|---|---|---|---|---|---|---|
| Becker et al. 2018 | P | 1 | 1 | 1 | 1 | 1 | 1 | 1 | 0 | 0 | U | 1 | 1 | 1 | 1 | 0 | 11 |
| Becker et al. 2017 | R | 1 | 1 | 1 | 1 | 1 | 1 | 0 | 0 | 0 | U | 1 | 1 | 1 | U | 0 | 9 |
| Loudon et al. 2012 | R | 1 | 1 | 1 | 1 | 1 | 1 | 1 | 0 | 0 | U | 1 | 1 | 1 | U | 0 | 10 |
| Schütte et al. 2018 | R | 1 | 1 | 1 | 1 | 1 | 1 | 1 | U | U | 0 | 1 | 1 | 1 | U | U | 10 |

#### Tibial stress fractures

| | Prospective (P) or retrospective (R) study | Clear aim/hypothesis | Outcome measures clearly described | Patient characteristics clearly described | Confounding variables described | Main findings clearly described | Measures of random variability provided | Actual probability values reported | Participants asked to participate representative of entire population | Participants prepared to participate representative of entire population | Blinding of outcome measurer | Analysis completed was planned | Appropriate statistics | Valid and reliable outcome measures | Appropriate case-control matching | Adjustment made for confounding variables | Total |
|---|---|---|---|---|---|---|---|---|---|---|---|---|---|---|---|---|---|
| Meardon et al. 2015 | R | 1 | 1 | 1 | 1 | 1 | 1 | 1 | 0 | 0 | U | 1 | 1 | 1 | 1 | 0 | 11 |
| Bennell et al. 2004 | R | 1 | 1 | 1 | 1 | 1 | 1 | 1 | 0 | 0 | U | 1 | 1 | 1 | 1 | 0 | 11 |
| Crossley et al. 1999 | R | 1 | 1 | 1 | 1 | 1 | 1 | 0 | 0 | 0 | U | 1 | 1 | 1 | 1 | 0 | 10 |
| Milner et al. 2006 | R | 1 | 1 | 1 | 0 | 1 | 1 | 1 | 0 | 0 | U | 1 | 1 | 1 | 1 | 0 | 10 |
| Milner et al. 2006 | R | 1 | 1 | 1 | 0 | 1 | 1 | 1 | 0 | 0 | U | 1 | 1 | 1 | 1 | 0 | 10 |
| Milner et al. 2010 | R | 1 | 1 | 1 | 1 | 1 | 1 | 1 | 0 | 0 | U | 1 | 1 | 1 | U | 0 | 10 |
| Pohl et al. 2008 | R | 1 | 1 | 1 | 1 | 1 | 1 | 0 | 0 | 0 | U | 1 | 1 | 1 | U | 0 | 9 |
| Milner et al. 2007 | R | 1 | 1 | 1 | 1 | 1 | 1 | 1 | 0 | 0 | U | 1 | 1 | 1 | U | 0 | 10 |
| Johnson et al., 2020 | R | 1 | 1 | 1 | 1 | 1 | 1 | 1 | U | U | 1 | 1 | 1 | 1 | U | U | 11 |

### Plantar fasciitis

| Black & Down Paper | Prospective (P) or retrospective (R) study | Clear aim/hypothesis | Outcome measures clearly described | Patient characteristics clearly described | Confounding variables described | Main findings clearly described | Measures of random variability provided | Actual probability values reported | Participants asked to participate representative of entire population | Participants prepared to participate representative of entire population | Blinding of outcome measurer | Analysis completed was planned | Appropriate statistics | Valid and reliable outcome measures | Appropriate case-control matching | Adjustment made for confounding variables | Total |
|---|---|---|---|---|---|---|---|---|---|---|---|---|---|---|---|---|---|
| Ribeiro et al. 2015 | R | 1 | 1 | 1 | 1 | 1 | 1 | 1 | U | U | U | U | 1 | 1 | 1 | 1 | 11 |
| Pohl et al. 2009 | R | 1 | 1 | 1 | U | 1 | 1 | 1 | 1 | 1 | 0 | U | 1 | 1 | U | U | 10 |
| Johnson et al. 2020 | R | 1 | 1 | 1 | 1 | 1 | 1 | 1 | U | U | 1 | 1 | 1 | 1 | U | U | 11 |
| Messier et al 1988 | R | 1 | 1 | 0 | U | 1 | U | 0 | U | U | 0 | 1 | 1 | 1 | U | U | 6 |
| Ribeiro et al. 2011 | R | 1 | 1 | 1 | 1 | 1 | U | 1 | 1 | U | 0 | 1 | 1 | 1 | U | U | 10 |

#### Patellofemoral pain syndrome

| | Prospective (P) or retrospective (R) study | Clear aim/hypothesis | Outcome measures clearly described | Patient characteristics clearly described | Confounding variables described | Main findings clearly described | Measures of random variability provided | Actual probability values reported | Participants asked to participate representative of entire population | Participants prepared to participate representative of entire population | Blinding of outcome measurer | Analysis completed was planned | Appropriate statistics | Valid and reliable outcome measures | Appropriate case-control matching | Adjustment made for confounding variables | Total |
|---|---|---|---|---|---|---|---|---|---|---|---|---|---|---|---|---|---|
| Luz et al., 2018 | R | 1 | 1 | 1 | 1 | 1 | 1 | 1 | U | U | 0 | 1 | 1 | 1 | 1 | 1 | 12 |
| Esculier et al. 2015 | R | 1 | 1 | 1 | 1 | 1 | 1 | 1 | U | U | 0 | 1 | 1 | 1 | 1 | 1 | 12 |
| Souza et al. 2009 | R | 1 | 1 | 0 | 1 | 1 | 1 | 1 | 1 | U | 0 | 1 | 1 | U | U | 1 | 10 |
| Souza et al. 2009 (PFPS6, same research group same sample) | R | 1 | 1 | 0 | 1 | 1 | 1 | 1 | 1 | U | 0 | 1 | 1 | U | U | 1 | 10 |
| Noehren et al. 2013 | P | 1 | 1 | 0 | 1 | 1 | U | 0 | 1 | 1 | 0 | 1 | 1 | 1 | 1 | 1 | 11 |
| Liao et al., 2018 | R | 1 | 1 | 1 | 1 | 1 | 1 | 1 | U | U | 0 | 1 | 1 | 1 | U | 1 | 11 |
| Eskofier et al. 2012 Stefanyshyn Data set | P | 0 | 0 | 1 | U | 1 | 1 | 0 | 1 | U | 0 | 0 | 1 | 1 | 1 | 1 | 8 |
| Willson et al. 2013 | R | 1 | 1 | 1 | 1 | 1 | 1 | 1 | U | U | 0 | 1 | 1 | 1 | 1 | 1 | 12 |
| Dierks et al. 2008 | R | 1 | 1 | 1 | 1 | 1 | 1 | 1 | 1 | U | 0 | U | 1 | 1 | 0 | 0 | 10 |
| Boldt et al., 2013 | R | 1 | 1 | 1 | 1 | 1 | 1 | 1 | U | U | 0 | 1 | 1 | 1 | 0 | 1 | 11 |
| Stepanyshyn et al. 2006 | P | 1 | 1 | 0 | 2 | 1 | 1 | 1 | U | U | U | U | 1 | 1 | 1 | 1 | 11 |
| Willson et al. 2011 | R | 1 | 1 | 1 | 2 | 1 | 0 | 1 | U | U | 0 | 1 | 1 | U | U | 1 | 10 |
| Willson et al. 2007 | R | 1 | 1 | 1 | 2 | 1 | 0 | 1 | U | U | 0 | 1 | 1 | U | U | 1 | 10 |
| Thijs et al. 2008 | P | 1 | 1 | 1 | 0 | 1 | 1 | 0 | U | U | 1 | 1 | 1 | 1 | 1 | 1 | 11 |
| Duffey et al. 2000 | R | 1 | 1 | 1 | 1 | 1 | 1 | 1 | U | U | 0 | 1 | 1 | 1 | U | U | 10 |
| Dierks et al. 2011 | R | 1 | 1 | 1 | 2 | 1 | 1 | 1 | U | U | 0 | 1 | 1 | 1 | 0 | 0 | 11 |
| Noehren et al. 2012 | R | 1 | 1 | 0 | U | 1 | U | 0 | 1 | 1 | 0 | 1 | 1 | 1 | 1 | 1 | 10 |

| | Prospective (P) or retrospective (R) study | Clear aim/hypothesis | Outcome measures clearly described | Patient characteristics clearly described | Confounding variables described | Main findings clearly described | Measures of random variability provided | Actual probability values reported | Participants asked to participate representative of entire population | Participants prepared to participate representative of entire population | Blinding of outcome measurer | Analysis completed was planned | Appropriate statistics | Valid and reliable outcome measures | Appropriate case-control matching | Adjustment made for confounding variables | Total |
|---|---|---|---|---|---|---|---|---|---|---|---|---|---|---|---|---|---|
| Rodrigues et al., 2013 | R | 1 | 1 | 1 | 1 | 1 | 1 | 1 | U | U | 0 | 1 | 1 | 1 | 0 | 1 | 11 |
| Johnson et al. 2020 | R | 1 | 1 | 1 | 1 | 1 | 1 | 1 | U | U | 0 | 1 | 1 | 1 | 1 | 1 | 12 |
| Messier et al., 1991 | R | 1 | 1 | 1 | 1 | 1 | 1 | 1 | U | U | 0 | 1 | 1 | 1 | 0 | 1 | 11 |
| Besier et al., 2009 | R | 1 | 1 | 0 | 1 | 1 | 1 | 1 | U | U | U | 1 | 1 | 1 | 0 | 1 | 10 |
| Bazett-Jones et al., 2013 | R | 1 | 1 | 1 | 1 | 1 | 1 | 1 | U | U | 0 | 1 | 1 | 1 | 1 | 1 | 12 |
| Cumningham et. al., 2014 | R | 1 | 1 | 1 | 1 | 1 | 1 | 1 | U | U | 0 | 1 | 1 | 1 | 0 | 1 | 11 |
| Liao et al., 2019 | R | 1 | 1 | 1 | 1 | 1 | 1 | 1 | U | U | 0 | 1 | 1 | 1 | U | 1 | 11 |
| Rees et al., 2018 | R | 1 | 1 | 1 | 1 | 1 | 1 | 1 | U | U | 0 | 1 | 1 | 1 | U | 1 | 11 |

#### Patellar tendinopathy

| | Prospective (P) or retrospective (R) study | Clear aim/hypothesis | Outcome measures clearly described | Patient characteristics clearly described | Confounding variables described | Main findings clearly described | Measures of random variability provided | Actual probability values reported | Participants asked to participate representative of entire population | Participants prepared to participate representative of entire population | Blinding of outcome measurer | Analysis completed was planned | Appropriate statistics | Valid and reliable outcome measures | Appropriate case-control matching | Adjustment made for confounding variables | Total |
|---|---|---|---|---|---|---|---|---|---|---|---|---|---|---|---|---|---|
| Grau et al., 2008 | R | 1 | 0 | 0 | 0 | 1 | 1 | 0 | U | U | U | 1 | 1 | 1 | 1 | 0 | 7 |

### Risk of Bias Assessment

#### Achilles tendinopathy

| | Definition of RROI | Study design<br>Pro = Yes<br>Retro = No | Description of runners or type of runners<br>Yes = e.g. recreational | Random sample selection process<br>Yes = Pro checking<br>No = Retro | Loss to follow-up<br>Yes = Pro & <20% drop out<br>No = not reported<br>N/A = Retro | Data collected directly from the runners always YES | Same mode of data collection always YES | Diagnosis conducted by any physicians<br>YES = if mentioned | Follow-up period<br>Yes = Pro & 6months+<br>Yes = Retro <12months | Incidence or prevalence rates of each RRMI expressed by any ratio | Score Risk of Bias |
|---|---|---|---|---|---|---|---|---|---|---|---|
| McCrory et al. 1999 | 0 | 0 | 1 | 0 | N/A | 1 | 1 | 1 | N/A | N/A | 4 |
| Donoghue et al. 2008 | 0 | 0 | 0 | 0 | N/A | 1 | 1 | 1 | 1 | N/A | 4 |
| Williams et al., 2008 | 0 | 0 | 1 | 0 | N/A | 1 | 1 | 1 | N/A | N/A | 4 |
| Azevedo et al., 2009 | 1 | 0 | 1 | 0 | N/A | 1 | 1 | 1 | N/A | N/A | 5 |
| Ryan et al. 2009 | 1 | 0 | 1 | 0 | N/A | 1 | 1 | 1 | 1 | N/A | 6 |
| van Ginckle et al. 2009 | 1 | 1 | 1 | 0 | 1 | 1 | 1 | 1 | 1 | N/A | 8 |
| Baur et al., 2011 | 1 | 0 | 0 | 0 | N/A | 1 | 1 | 1 | N/A | N/A | 4 |
| Wyndow et al. 2013 | 1 | 0 | 1 | 0 | N/A | 1 | 1 | N/A | 1 | N/A | 5 |
| Franettovich et al. 2014 | 1 | 0 | 1 | 0 | N/A | 1 | 1 | N/A | N/A | N/A | 4 |
| Creaby et al. 2016 | 1 | 0 | 1 | 0 | N/A | 1 | 1 | N/A | N/A | N/A | 4 |
| Becker et al. 2017 | 0 | 0 | 1 | 0 | N/A | 1 | 1 | 1 | 0 | N/A | 4 |
| Johnson et al. 2020 | 0 | 0 | 0 | 0 | N/A | 1 | 1 | 1 | 0 | N/A | 3 |

#### Iliotibial band syndrome

| | Definition of RROI | Study design<br>Pro = Yes<br>Retro = No | Description of runners or type of runners<br>Yes = e.g. recreational | Random sample selection process<br>Yes = Pro checking<br>No = Retro | Loss to follow-up<br>Yes = Pro & <20% drop out<br>No = not reported<br>N/A = Retro | Data collected directly from the runners always YES | Same mode of data collection always YES | Diagnosis conducted by any physicians<br>YES = if mentioned | Follow-up period<br>Yes = Pro & 6months+<br>Yes = Retro <12months | Incidence or prevalence rates of each RRMI expressed by any ratio | Score Risk of Bias |
|---|---|---|---|---|---|---|---|---|---|---|---|
| Brown et al. 2019 | 0 | 0 | 1 | 0 | N/A | 1 | 1 | 1 | 1 | N/A | 5 |
| Baker et al. 2018 | 1 | 0 | 1 | 0 | N/A | 1 | 1 | 1 | 1 | N/A | 6 |
| Brown et al. 2016 | 0 | 0 | 1 | 0 | N/A | 1 | 1 | 1 | 1 | N/A | 5 |
| Foch et al. 2015 | 1 | 0 | 1 | 0 | N/A | 1 | 1 | 1 | 0 | N/A | 5 |
| Phinyomark et al., 2014 | 1 | 0 | 1 | 0 | N/A | 1 | 1 | 1 | 1 | N/A | 6 |
| Noehren et al. 2014 | 1 | 0 | 1 | 0 | N/A | 1 | 1 | 1 | 1 | N/A | 6 |
| Foch and Milner 2014 | 1 | 0 | 1 | 0 | N/A | 1 | 1 | 1 | N/A | N/A | 5 |
| Ferber et al. 2010 | 0 | 0 | 1 | 0 | N/A | 1 | 1 | 1 | N/A | N/A | 4 |
| Grau et al 2008 | 0 | 0 | 1 | 0 | N/A | 1 | 1 | 1 | N/A | N/A | 4 |
| Hamill et al. 2008 | 0 | 1 | 1 | 0 | N/A | 1 | 1 | 1 | N/A | N/A | 5 |
| Noehren et al. 2007 | 0 | 1 | 1 | 0 | N/A | 1 | 1 | 1 | N/A | N/A | 5 |
| Messier et al. 1995 | 1 | 0 | 1 | 0 | N/A | 1 | 1 | 1 | N/A | N/A | 5 |
| Johnson et al., 2020 | 0 | 0 | 0 | 0 | N/A | 1 | 1 | 1 | 0 | N/A | 3 |
| Grau et al 2011 | 1 | 0 | 1 | 0 | N/A | 1 | 1 | 1 | N/A | N/A | 5 |
| Messier et al 1988 | 1 | 0 | 1 | 0 | N/A | 1 | 1 | 1 | 0 | N(A | 5 |
| Millert et al. 2007 | 1 | 0 | 1 | 0 | N/A | 1 | 1 | 1 | U | N/A | 5 |

### Medial tibial stress syndrome

| | Definition of RROI | Study design<br>Pro = Yes<br>Retro = No | Description of runners or<br>type of runners<br>Yes = e.g. recreational | Random sample selection process<br>Yes = Pro checking<br>No = Retro | Loss to follow-up<br>Yes = Pro & <20% drop out<br>No = not reported<br>N/A = Retro | Data collected directly from the runners always YES | Same mode of data collection always YES | Diagnosis conducted by<br>any physicians<br>YES = if mentioned | Follow-up period<br>Yes = Pro & 6months+<br>Yes = Retro <12months | Incidence or prevalence<br>rates of each RRMI expressed by any ratio | Score Risk of Bias |
|---|---|---|---|---|---|---|---|---|---|---|---|
| **Becker et al. 2018** | **0** | **1** | **1** | **1** | **1** | **1** | **1** | **0** | **1** | **N/A** | 7 |
| **Becker et al. 2017** | **0** | **0** | **1** | **0** | **N/A** | **1** | **1** | **1** | **N/A** | **N/A** | 4 |
| **Loudon et al. 2012** | **0** | **0** | **1** | **0** | **N/A** | **1** | **1** | **1** | **N/A** | **N/A** | 4 |
| **Schütte et al. 2018** | **1** | **0** | **1** | **0** | **N/A** | **1** | **1** | **0** | **1** | **N/A** | 5 |

### Tibial stress fractures

| Risk of Bias | Definition of RROI | Study design<br>Pro = Yes<br>Retro = No | Description of runners or<br>type of runners<br>Yes = e.g. recreational | Random sample selection process<br>Yes = Pro checking<br>No = Retro | Loss to follow-up<br>Yes = Pro & <20% drop out<br>No = not reported<br>N/A = Retro | Data collected directly from the runners always YES | Same mode of data collection always YES | Diagnosis conducted by<br>any physicians<br>YES = if mentioned | Follow-up period<br>Yes = Pro & 6months+<br>Yes = Retro <12months | Incidence or prevalence<br>rates of each RRMI expressed by any ratio | Score Risk of Bias |
|---|---|---|---|---|---|---|---|---|---|---|---|
| **Meardon et al. 2015** | **0** | **0** | **1** | **0** | **N/A** | **1** | **1** | **1** | **N/A** | **N/A** | 4 |
| **Bennell et al. 2004** | **0** | **0** | **1** | **0** | **N/A** | **1** | **1** | **1** | **N/A** | **N/A** | 4 |
| **Crossley et al. 1999** | **0** | **0** | **1** | **0** | **N/A** | **1** | **1** | **1** | **N/A** | **N/A** | 4 |
| **Milner et al. 2006** | **0** | **0** | **1** | **0** | **N/A** | **1** | **1** | **1** | **0** | **N/A** | 4 |
| **Milner et al. 2006** | **0** | **0** | **1** | **0** | **N/A** | **1** | **1** | **1** | **0** | **N/A** | 4 |
| **Milner et al. 2010** | **1** | **0** | **1** | **0** | **N/A** | **1** | **1** | **1** | **N/A** | **N/A** | 5 |
| **Pohl et al. 2008** | **0** | **0** | **1** | **0** | **N/A** | **1** | **1** | **1** | **N/A** | **N/A** | 4 |
| **Milner et al. 2007** | **0** | **0** | **1** | **0** | **N/A** | **1** | **1** | **1** | **N/A** | **N/A** | 4 |
| **Johnson et al., 2020** | **0** | **0** | **0** | **0** | **N/A** | **1** | **1** | **1** | **0** | **N/A** | 3 |

#### Plantar fasciitis

| | Definition of RROI | Study design<br>Pro = Yes<br>Retro = No | Description of runners or type of runners<br>Yes = e.g. recreational | Random sample selection process<br>Yes = Pro checking<br>No = Retro | Loss to follow-up<br>Yes = Pro & <20% drop out<br>No = not reported<br>N/A = Retro | Data collected directly from the runners always YES | Same mode of data collection always YES | Diagnosis conducted by any physicians<br>YES = if mentioned | Follow-up period<br>Yes = Pro & 6months+<br>Yes = Retro <12months | Incidence or prevalence rates of each RRMI expressed by any ratio | Score Risk of Bias |
|---|---|---|---|---|---|---|---|---|---|---|---|
| Ribeiro et al. 2015 | 1 | 0 | 1 | 0 | N/A | 1 | 1 | 1 | 0 | N/A | 5 |
| Pohl et al. 2009 | 0 | 0 | 1 | 0 | N/A | 1 | 1 | 1 | 0 | N/A | 4 |
| Johnson et al. 2020 | 0 | 0 | 0 | 0 | N/A | 1 | 1 | 1 | 0 | N/A | 3 |
| Messier et al 1988 | 1 | 0 | 1 | 0 | N/A | 1 | 1 | 1 | 0 | N(A | 5 |
| Ribeiro et al. 2011 | 1 | 0 | 1 | 0 | N/A | 1 | 1 | 1 | 0 | N/A | 5 |

#### Patellofemoral pain syndrome

| | Definition of RROI | Study design<br>Pro = Yes<br>Retro = No | Description of runners or type of runners<br>Yes = e.g. recreational | Random sample selection process<br>Yes = Pro checking<br>No = Retro | Loss to follow-up<br>Yes = Pro & <20% drop out<br>No = not reported<br>N/A = Retro | Data collected directly from the runners always YES | Same mode of data collection always YES | Diagnosis conducted by any physicians<br>YES = if mentioned | Follow-up period<br>Yes = Pro & 6months+<br>Yes = Retro <12months | Incidence or prevalence rates of each RRMI expressed by any ratio | Score Risk of Bias |
|---|---|---|---|---|---|---|---|---|---|---|---|
| Luz et al., 2018 | 1 | 0 | 1 | 0 | N/A | 1 | 1 | 0 | 1 | N/A | 5 |
| Esculier et al. 2015 | 1 | 0 | 1 | 0 | N/A | 1 | 1 | 0 | 1 | N/A | 5 |
| Souza et al. 2009 | 1 | 0 | 0 | 0 | N/A | 1 | 1 | 1 | N/A | N/A | 4 |
| Noehren et al. 2013 | 1 | 1 | 1 | 0 | 1 | 1 | 1 | 1 | 1 | N/A | 8 |
| Liao et al., 2018 | 1 | 0 | 1 | 0 | N/A | 1 | 1 | 0 | N/A | N/A | 4 |
| Eskofier et al. 2012t | 1 | 1 | 1 | 0 | 0 | 1 | 1 | 1 | 1 | N/A | 7 |
| Willson et al. 2013 | 1 | 0 | 1 | 0 | N/A | 1 | 1 | 1 | 1 | N/A | 6 |
| Dierks et al. 2008 | 1 | 0 | 1 | 0 | 0 | 1 | 1 | 1 | N/A | N/A | 5 |
| Boldt et al., 2013 | 1 | 0 | 1 | 0 | N/A | 1 | 1 | 1 | N/A | N/A | 5 |
| Stepanyshyn et al. 2006 | 1 | 1 | 1 | 0 | 0 | 1 | 1 | 1 | 1 | N/A | 7 |
| Willson et al. 2011 | 1 | 0 | 1 | 0 | N/A | 1 | 1 | 1 | N/A | N/A | 5 |
| Willson et al. 2007 | 1 | 0 | 1 | 0 | N/A | 1 | 1 | 1 | N/A | N/A | 5 |
| Thijs et al. 2008 | 1 | 1 | 0 | 0 | 1 | 1 | 1 | 1 | 1 | N/A | 7 |
| Duffey et al. 2000 | 1 | 0 | 1 | 0 | N/A | 1 | 1 | 1 | N/A | N/A | 5 |
| Dierks et al. 20011 | 1 | 0 | 1 | 0 | N/A | 1 | 1 | 1 | 1 | N/A | 6 |
| Noehren et al. 2012 | 1 | 0 | 1 | 0 | N/A | 1 | 1 | 1 | 1 | N/A | 6 |
| Rodrigues et al., 2013 | 0 | 0 | 0 | 0 | N/A | 1 | 1 | 1 | 1 | N/A | 4 |
| Johnson et al. 2020 | 0 | 0 | 1 | 0 | N/A | 1 | 1 | 1 | 1 | N/A | 5 |
| Messier et al., 1991 | 1 | 0 | 1 | 0 | N/A | 1 | 1 | 1 | 1 | N/A | 6 |
| Besier et al., 2009 | 1 | 0 | 0 | 0 | N/A | 1 | 1 | 1 | N/A | N/A | 4 |

| | Definition of RROI | Study design<br>Pro = Yes<br>Retro = No | Description of runners or type of runners<br>Yes = e.g. recreational | Random sample selection process<br>Yes = Pro checking<br>No = Retro | Loss to follow-up<br>Yes = Pro & <20% drop out<br>No = not reported<br>N/A = Retro | Data collected directly from the runners always YES | Same mode of data collection always YES | Diagnosis conducted by any physicians<br>YES = if mentioned | Follow-up period<br>Yes = Pro & 6months+<br>Yes = Retro <12months | Incidence or prevalence rates of each RRMI expressed by any ratio | Score Risk of Bias |
|---|---|---|---|---|---|---|---|---|---|---|---|
| Cumningham et. al., 2014 | 0 | 0 | 1 | 0 | N/A | 1 | 1 | 1 | U | N/A | 4 |
| Liao et al., 2019 | 1 | 0 | 1 | 0 | N/A | 1 | 1 | 0 | 1 | N/A | 5 |
| Rees et al., 2018 | 1 | 0 | 1 | 0 | N/A | 1 | 1 | 1 | 1 | N/A | 6 |

#### Patellar tendinopathy

| | Definition of RROI | Study design<br>Pro = Yes<br>Retro = No | Description of runners or type of runners<br>Yes = e.g. recreational | Random sample selection process<br>Yes = Pro checking<br>No = Retro | Loss to follow-up<br>Yes = Pro & <20% drop out<br>No = not reported<br>N/A = Retro | Data collected directly from the runners always YES | Same mode of data collection always YES | Diagnosis conducted by any physicians<br>YES = if mentioned | Follow-up period<br>Yes = Pro & 6months+<br>Yes = Retro <12months | Incidence or prevalence rates of each RRMI expressed by any ratio | Score Risk of Bias |
|---|---|---|---|---|---|---|---|---|---|---|---|
| Grau et al., 2008 | 0 | 0 | U | 0 | N/A | 1 | 1 | 1 | U | N/A | 3 |
