## Supplementary Digital Content 3 for "Running related biomechanical risk factors for overuse injuries in distance runners: A systematic review considering injury specificity and the potentials for future research"

| Author | Injury | Study Design | Population | Gender | Protocol |
|---|---|---|---|---|---|
| Messier et al. 1991 | PFPS | R | N=20 healthy, N=16 injured | mixed | 2D Video Kinematics. GRFs |
| Duffey et al. 2000 | PFPS | R | N=70 healthy, N=99 injured | mixed | 3D Kinematics, Kinetics |
| Stefanyshyn et al. 2006 | PFPS | P | N=80 healthy, N=6 injured | mixed | 3D Kinematics, Kinetics |
| Willson et al.2008 | PFPS | R | N=20 healthy, N=20 injured | females | 3D Kinematics, Kinetics |
| Thijs et al. 2008 | PFPS | P | N=102 healthy, N=17 injured | mixed | Plantar pressure |
| Dierks et al. 2008 | PFPS | R | N=20 heathly, N=20 injured | females | 3D Kinematics |
| Souza et al. 2009 | PFPS | R | N=20 healthy, N=21 injured | females | 3D Kinematics, GRFs, EMG |
| Besier et al. 2009 | PFPS | R | N=27 healthy, N=16 injured | mixed | 3D Kinematics, Kinetics, EMG |
| Willson et al. 2011 | PFPS | R | N=20 healthy, N=20 injured | females | 3D Kinematics, GRFs, EMG |
| Noehren et al. 2012 | PFPS | R | N=16 healthy, N=16 injured | females | 3D Kinematics, GRFs |

| | | | | | |
|---|---|---|---|---|---|
| Eskofier et al. 2012 | PFPS | P | N=80 healthy, N=6 injured | mixed | 3D Kinematics, Kinetics |
| Rodrigues et al. 2013 | PFPS | R | N=19 healthy, N=17 injured | mixed | 3D Kinematics |
| Boldt et al. 2013 | PFPS | R | N=19 healthy, N=19 injured | females | 3D Kinematics |
| Noehren et al. 2013 | PFPS | P | N=400 healthy, N=15 injured | females | 3D Kinematics, GRFs |
| Bazett-Jones et al. 2013 | PFPS | R | N=19 healthy, N=19 injured | mixed | 3D Kinematics, Kinetics |
| Willson et al. 2013 | PFPS | R | N=13 healthy, N=10 injured | females | 3D Kinematics, Kinetics |
| Cumningham et al. 2014 | PFPS | R | N=11 healthy, N=19 injured | mixed | 3D Kinematics, GRFs |
| Escilier et al. 2015 | PFPS | R | N=20 healthy, N=21 injured | mixed | 3D Kinematics, GRFs, EMG |
| Rees et al. 2018 | PFPS | R | N=16 healthy, N=16 injured | mixed | 2D Video Kinematics |
| Liao et al. 2018 | PFPS | R | N=10 healthy, N=12 injured | females | 3D Kinematics, Kinetics, EMG |
| Luz et al. 2018 | PFPS | R | N=27 healthy, N=27 injured | mixed | 3D Kinematics |
| Liao et al. 2019 | PFPS | R | N=10 healthy, N=12 injured | females | 3D Kinematics, Kinetics, EMG |
| Johnson et al. 2020 | PFPS | R | N=65 healthy, N=31 injured | mixed | GRFs |
| Grau et al. 2008 | PT | R | N=12 healthy, N=12 injured | females | 3D Kinematics |
| Messier et al. 1988 | PF | R | N=19 healthy, N=15 injured | mixed | 2D Video Kinematics |
| Pohl et al. 2009 | PF | R | N=25 healthy, N=25 injured | females | 3D Kinematics, GRFs |
| Ribeiro et al. 2011 | PF | R | N=60 healthy, N=30 current injured, N=15 previous injured | mixed | Pressure insoles |

| | | | | | |
|---|---|---|---|---|---|
| Ribeiro et al. 2015 | PF | R | N=30 healthy, N=30 acute, N=30 chronic | mixed | Pressure insoles |
| Johnson et al. 2020 | PF | R | N=65 healthy, N=22 injured | mixed | GRFs |
| Crossley et al. 1999 | TSF | R | N=23 healthy, N=23 injured | males | GRFs |
| Bennell et al. 2004 | TSF | R | N=23 healthy, N=13 injured | females | GRFs |
| Milner et al. 2005 | TSF | R | N=25 healthy, N=25 injured | females | 3D Kinematics, GRFs |
| Milner et al. 2006 | TSF | R | N=20 healthy, N=20 injured | females | 3D Kinematics, Kinetics |
| Milner et al. 2007 | TSF | R | N=23 healthy, N=23 injured | females | 3D Kinematics, Kinetics |
| Pohl et al. 2008 | TSF | R | N=30 healthy, N=30 injured | females | 3D Kinematics, GRFs |
| Milner et al. 2010 | TSF | R | N=30 healthy, N=30 injured | females | 3D Kinematics, GRFs |
| Meardon et al. 2015 | TSF | R | N=23 healthy, N=23 injured | mixed | 3D Kinematics, Kinetics |
| Johnson et al. 2020 | TSF | R | N=65 healthy, N=23 injured | mixed | GRFs |
| Brown et al. 2019 | ITBS | R | N = 20 healthy, N = 12 current injured | females | 3D Kinematics, EMG |
| Baker et al. 2018 | ITBS | R | N = 15 healthy, N= 15 injured | mixed | 3D Kinematics, EMG |
| Brown et al. 2016 | ITBS | R | N = 20 healthy, N= 12 injured | females | 3D Kinematics, Kinetics |
| Foch et al. 2015 | ITBS | R | N = 9 healthy, N= 9 current injured, N = 9 previous injured | females | 3D Kinematics, Kinetics |
| Noehren et al. 2014 | ITBS | R | N = 17 healthy, N= 17 injured | males | 3D Kinematics, GRFs |
| Foch and Milner 2014 | ITBS | R | N = 20 healthy, N= 20 injured | females | 3D Kinematics, Kinetics |

| | | | | | |
|---|---|---|---|---|---|
| Ferber et al. 2010 | ITBS | R | N = 35 healthy, N= 35 injured | females | 3D Kinematics, Kinetics |
| Grau et al 2008 | ITBS | R | N = 52 healthy, N= 18 injured | mixed | 3D Kinematics, plantar pressure |
| Hamill et al. 2008 | ITBS | P | N = 17 healthy, N= 17 injured | females | 3D Kinematics, GRFs |
| Noehren et al. 2007 | ITBS | P | N = 18 healthy, N= 18 injured | females | 3D Kinematics, Kinetics |
| Messier et al. 1995 | ITBS | R | N = 70 healthy, N= 56 injured | mixed | 2D Kinematics, GRFs |
| Johnson et al. 2020 | ITBS | R | N = 65 healthy, N= 28 injured | mixed | GRFs |
| Grau et al 2011 | ITBS | R | N = 18 healthy, N= 18 injured | mixed | 3D Kinematics |
| Messier et al. 1988 | ITBS | R | N = 19 healthy, N= 13 injured | mixed | 2D Kinematics |
| Loudon et al. 2012 | MTSS | R | N = 14 healthy, N= 14 injured | mixed | 3D Kinematics |
| Becker et al. 2017 | MTSS | R | N = 21 healthy, N= 8 injured | mixed | 3D Kinematics, GRFs |
| Becker et al. 2018 | MTSS | P | N = 24 healthy, N= 7 injured | mixed | 3D Kinematics |
| Schütte et al. 2018 | MTSS | R | N = 16 healthy, N= 14 injured | mixed | Accelerometer |
| McCrory et al. 1999 | AT | R | N = 58 healthy, N = 31 injured | mixed | Dynamometer, 2D Kinematics, Kinetics |
| Donoghue et al. 2008 | AT | R | N = 12 healthy, N = 12 injured | mixed | 3D Kinematics |
| Williams et al. 2008 | AT | R | N = 8 injured, N = 8 healthy | mixed | 3D Kinematics, Kinetics |
| Azevedo et al. 2009 | AT | R | N=21 healthy N = 21 injured | mixed | 3D Kinematics, GRFs, EMG |
| Ryan et al. 2009 | AT | R | N = 27 healthy, N = 21 injured | males | 3D Kinematics |

| | | | | | |
|---|---|---|---|---|---|
| van Ginckle et al. 2009 | AT | P | N = 129, N = 10 injured (3 bilateral), excluded = 66, N total = 63 | mixed | Pressure Plate |
| Baur et al 2011 | AT | R | N = 30 healthy, N = 30 injured | mixed | EMG |
| Wyndow et al. 2013 | AT | R | N=15 injured, N =19 healthy | males | EMG, GRFs |
| Franettovich et al. 2014 | AT | R | N = 19 healthy, N = 14 injured | males | EMG, GRFs |
| Creaby et al. 2017 | AT | R | N=11 healthy, N = 14 njured | males | 3D Kinematics, Kinetics |
| Becker et al. 2017 | AT | R | N = 21 healthy, N = 13 AT injured, N = 8 injured | mixed | 3D Kinematics, GRFs |
| Johnson et al. 2020 | AT | R | N = 65 healthy, N = 21 injured | mixed | GRFs |

**erview**
**uries in distance runners:**
**otentials for future research**

Joseph Hamill[4], Luke Kelly[5], Patrick Mai[3]

Applied Sciences, Offenburg, Germany
iversity, Östersund, Sweden
e, Cologne, Germany

, St. Lucia, Queensland, Australia

| Setting | Running Speed | Footwear used (O= own, C = controlled labratory shoe, B = Bare, N/A=not applicable |
|---|---|---|
| Overground | self selected | N/A |
| Treadmill | self selected | O |
| Overground | 4.0 ± 0.2 m/s | O |
| Overground | 3.7 ± 0.11 m/s | C |
| Overground | self selected | B |
| Treadmill | self selected | C |
| Overground | 3 ± 0.15 m/s | N/A |
| Overground | self selected | N/A |
| Overground | 3.52 - 3.83 m/s | C |
| Treadmill | 3.3 m/s | C |

| | | |
|---|---|---|
| Overground | 4.0 ± 0.2 m/s | O |
| Treadmill | 2.9 m/s | C |
| Overground | 3.52 - 3.83 m/s | C |
| Overground | 3.7 ± 0.18 m/s | C |
| Overground | self selected | N/A |
| Overground | 3.7 ± 0.18 m/s | C |
| Treadmill | self selected | O |
| Treadmill | self selected | O |
| Treadmill | self selected | O |
| Overground | 2.7 ± 0.27 m/s | N/A |
| Treadmill | self selected | C |
| Overground | 2.7 ± 0.27 m/s | N/A |
| Treadmill | self selected | C |
| Overground | 3.3 ± 0.17 m/s | B |
| Treadmill | self selected | O |
| Overground | 3.7 ± 0.19 m/s | C |
| Overground | 3.33 ± 0.17 m/s | C |

| | | |
|---|---|---|
| Overground | 3.33 ± 0.17 m/s | C |
| Treadmill | self selected | C |
| Overground | 4 ± 0.4 m/s | N/A |
| Overground | 5 ± 0.4 m/s | O |
| Overground | 3.7 ± 0.19 m/s | C |
| Overground | 3.7 ± 0.19 m/s | C |
| Overground | 3.7 ± 0.19 m/s | C |
| Overground | 3.7 ± 0.19 m/s | C |
| Overground | 3.7 ± 0.19 m/s | C |
| Overground | 3.7 m/s | C |
| Treadmill | self selected | C |
| Overground/ Treadmill | self selected | C |
| Treadmill | 2.74 m/s | O |
| Treadmill | 3.35 ± 0.34 m/s | C |
| Overground | 3.5 ± 0.17 m/s | N/A |
| Treadmill | 3.3 m/s | C |
| Overground | 3.5 ± 0.18 m/s | C |

| | | |
|---|---|---|
| Overground | 3.65 ± 0.1825 m/s | C |
| Overground | 3.3 ± 0.17 m/s | B |
| Overground | 3.7 ± 0.19 m/s | C |
| Overground | 3.7 ± 0.19 m/s | C |
| Overground/ Treadmill | self selected | O |
| Treadmill | self selected | O |
| Overground | 3.3 m/s | B |
| Treadmill | self selected | O |
| Treadmill | self selected | O |
| Overground | self selected | O |
| Treadmill | self selected | C |
| Overground | self selected | O |
| Overground | self selected | O |
| Treadmill | self selected | O |
| Overground | 3.35 ± 0.17 m/s | N/A |
| Overground | self selected | C |
| Overground | self selected | B |

| | | |
|---|---|---|
| Overground | self selected | B |
| Treadmill | 3.33 m/s | C |
| Overground | 4.00 m/s ± 0.1 m/s | C |
| Overground | 4.00 m/s ± 0.4 m/s | C |
| Overground | 4.00 m/s ± 0.4 m/s | C |
| Overground | self selected | O |
| Treadmill | self selected | C |
